# Perimenopause, DAN, and Cognition: An fMRI Study

**DOI:** 10.1101/2025.03.20.25324344

**Authors:** Ningning Liu, Yue Zhang, Weiqing Fu, Huijun Liu

**Author notes:** Correspondence: Ningning Liu, Department of Ultrasound, Second Hospital of Tianjin Medical University, Tianjin 300211, China, Address: Hexi pingjiang road NO.23, Tianjin, China.

## Abstract

**Objective:** To evaluate the functional changes of the dorsal attention network (DAN) in perimenopausal women using functional magnetic resonance imaging (fMRI) and explore the relationship between sex hormones and cognitive function.

**Methods:** A total of 25 perimenopausal women and 25 premenopausal women underwent sex hormone level measurements, scale - based assessments, cognition evaluations, and magnetic resonance imaging (MRI) scans. Resting - state fMRI data were acquired using a 3.0 Tesla magnetic resonance scanner. Independent component analysis (ICA) was employed to assess the differences in DAN functional connections between the two groups. Gray matter volume (GMV) values of brain regions with differences in DAN functional connections were extracted, and the GMV differences between the two groups were compared. Correlation analyses were performed between the connection strengths of DAN functional connections, GMV values of regions of interest (ROIs), sex hormone levels, and clinical and neuropsychological assessment results in both groups. Additionally, sensitivity and exploratory analyses were conducted on the existing data, and the results were compared with those of existing studies.

**Results:** Compared with the premenopausal group, the perimenopausal group showed enhanced functional connections in the right inferior parietal lobule (IPL) and the right angular gyrus (AG) within the DAN. There were no significant differences in GMV values between the two groups. Correlation analysis indicated that in perimenopausal women, the connection strength of the right IPL negatively correlated with the estradiol level and positively correlated with the reaction time of the STROOP color - word test. Sensitivity analysis showed that the main results were robust to the influence of extreme values. Exploratory analysis identified potential subgroups within perimenopausal women. Comparison with existing studies confirmed the consistency of some findings while also revealing differences.

**Conclusions:** ICA results suggest that DAN functional changes in perimenopausal women may trigger the brain’s compensatory mechanisms to cope with physiological and psychological challenges during the reproductive transition. These findings contribute to a better understanding of the brain function changes in perimenopausal women. However, limitations such as a small sample size still exist, and future research directions are proposed.

## Introduction

The perimenopausal period is a crucial transition in a woman’s life, marking the gradual decline of reproductive function. It is estimated that nearly two - thirds of women experience various symptoms during this stage, with cognitive - related complaints being common^1^. For example, studies have reported that perimenopausal women often struggle with concentration, experience forgetfulness, and perceive difficulties in cognitive tasks^2–4^.

Previous research has shown that cognitive function in perimenopausal women may be affected by hormonal changes, particularly the decline in estrogen levels^5–7^. Estrogen plays a vital role in the brain, influencing neural plasticity, neurotransmitter systems, and cerebral blood flow^8–10^. A decrease in estrogen during perimenopause has been associated with reduced metabolism in brain regions involved in learning and memory, such as the hippocampus and parahippocampal gyrus^11–12^.

The dorsal attention network (DAN), which consists of the bilateral parietal and frontal lobes, is essential for cognitive processing. It is responsible for top - down attentional control and exogenous attentional orientation^13–14^. However, the impact of perimenopause on the DAN remains unclear. Understanding these changes is crucial for developing targeted interventions to improve the cognitive health of perimenopausal women.

Functional magnetic resonance imaging (fMRI) has emerged as a powerful tool in neuroscience research. It allows for the non - invasive investigation of brain function and connectivity. In the context of perimenopause, fMRI can help us understand how hormonal fluctuations affect the brain’s functional architecture^15–17^.

In this study, we aimed to use fMRI and ICA to identify differences in the DAN between premenopausal and perimenopausal women. We also explored the relationship between serum hormone levels and DAN functional connectivity in perimenopausal women and evaluated the associations among DAN functional connectivity, serum estrogen levels, and cognitive function. Additionally, we conducted in - depth analysis of the existing data and compared our results with those of previous studies to enhance the reliability and comprehensiveness of our findings.

## Materials and Methods

### Participants

The recruitment period for this study started on March 1, 2024, and ended on October 1, 2024. The inclusion criteria were as follows: (a) female participants aged 45 - 55 years; (b) right - handedness; (c) having received more than 12 years of education; and (d) a diagnosis of perimenopause based on the Stages of Reproductive Aging Workshop (STRAW)+10 criteria (menstrual cycle change > 7 days, repeated for 10 menstrual cycles or amenorrhea interval ≥ 60 days, and follicle - stimulating hormone (FSH) range 11 - 45 IU/L). Premenopausal women were recruited if they did not meet the STRAW+10 diagnosis criteria, had a regular ovulation day determined by the rhythm method, and an FSH level < 11 IU/L.

The exclusion criteria included: (a) a history of neoplasms in the female genital organs, uterectomy, or oophorectomy; (b) the presence of neurological and psychiatric disorders, a history of brain trauma, smoking or alcohol dependence, or other diseases that could affect brain structure and function; (c) a diagnosis of mood disorders (such as depression or anxiety disorders); (d) a history of hormone administration; (e) color - blindness; or (f) MRI contraindications.

After screening, 50 participants were enrolled in the fMRI experiment, with 25 in the perimenopausal group (average age: 53.19±3.82 years) and 25 in the premenopausal group (average age: 47.67±3.48 years). All participants provided written informed consent, which was obtained before the start of any study - related procedures. The informed consent form was clearly explained to the participants, including the study purpose, procedures, potential risks and benefits.

### Sex Hormone Level Measurement

All participants had their sex hormone levels measured, including follicle - stimulating hormone (FSH), luteinizing hormone (LH), estradiol (E2), progesterone (P), testosterone (T), and prolactin. Blood samples were collected from the elbow vein between 8:00 - 9:00 am within 3 days of the start of menstruation. For participants with an abnormal menstrual cycle or amenorrhea, the blood collection was done at 8:00 - 9:00 am on the day of the experiment. The concentrations of the six sex hormones were determined using chemiluminescence analysis.

### Scale and Cognition Evaluations

The Menopause Rating Scale (MRS)^18^ was used to assess menopausal symptoms, and the Patient Health Questionnaire (PHQ - 9)^19–20^ was employed to screen for depressive symptoms. All participants also completed the computer - based STROOP color - word test. In this test, incongruent color words were presented in the center of a computer monitor, and participants were required to select the correct matching color word based on the ink color of the central word.

### fMRI Acquisition and Processing

All participants underwent conventional MRI examinations and resting - state fMRI scans using a 3.0 T MR scanner (Discovery MR 750, GE, US) with an eight - channel head coil. For participants with regular menstrual cycles, the fMRI scan was scheduled 3 days after the menstrual period. For those with amenorrhea, there was no such time restriction.

A three - dimensional brain volume (T1 3D - BRAVO) sequence was used for MRI scanning (TR/TE = 8.1/3.1 ms, flip angle = 13°, FOV = 256 mm × 256 mm, matrix = 256 × 256, slice = 176, slice thickness = 1 mm) to exclude organic lesions. The resting - state fMRI scans were acquired using a single - shot gradient echo planner imaging sequence (TR/TE = 2000/30 ms, FOV = 220 mm × 220 mm, matrix = 64 × 64, slice = 32, slice thickness = 3 mm, slice gap = 0.9 mm, and measurements = 180).

Before the scan, participants’ heads were stabilized with sponge pads, and they were instructed to keep their eyes closed, remain quiet, and stay awake. The fMRI data were analyzed using the Data Processing Assistant for Resting - State fMRI (DPARSFA, http://restfmri.net/forum/DPARSF) software. The first 10 volumes of the acquired images were discarded to account for BOLD signal stabilization and participant adaptation. The remaining 170 - volume data were preprocessed, including correction for acquisition time differences between layers and head movement. Images with translation ≥ 2 mm and rotation ≥ 2° were excluded. All images were then normalized to the Montreal Neurological Institute (MNI) template, and the data were resampled to obtain functional image data with 2 × 2 × 2 mm³ voxels. A Gaussian kernel with a full - width at half - maximum (FWHM) of 4×4×4 mm³ was used for spatial smoothing.

### Identification of Resting - State Networks (RSNs)

Independent component analysis (ICA) of the smoothed data was performed using MICA software tools (Stable and Consistent Group ICA of fMRI Toolbox, version1.2, http://www.nitrc.org/projects/cogicat/) based on the MatlabR2012a (MathWorks Inc., http://www.mathworks.com) platform. Data reduction was achieved through triple principal component analyses. The data for each participant were segmented into 20 spatially independent components, and the ICA arithmetic operation was repeated 100 times. After that, the independent components consistent with the literature - reported template were selected.

### Structural MRI Preprocessing

Structural MRI data were preprocessed using voxel - based morphometry and the Statistical Parametric Mapping (SPM12, London, UK) software. The data were segmented into gray matter (GM), white matter, and cerebrospinal fluid, and the GM DARTEL template was registered to the tissue probability map in the MNI space. Each voxel’s gray matter volume (GMV) was calculated by multiplying the GM concentration map by the nonlinear determinants from the spatial normalization step. The GMV maps were then smoothed with a Gaussian kernel of 4×4×4 mm³ FWHM.

### Statistical Analysis

Statistical analyses were carried out using SPSS 20.0 software (SPSS, Chicago, IL). The independent two - sample t - test was used to compare the age, years of education, MRS score, PHQ - 9 score, the accuracy rate and reaction time of the Color - Word STROOP, and sex hormone levels between the two groups. A significance level of P<0.05 was set.

The statistical analysis module of Matlab - based statistical parametric mapping (SPM12) was used to analyze RSNs. A general linear model was applied to detect significant differences in RSNs between premenopausal and perimenopausal women, with age and years of education controlled as covariates. For multiple comparison correction, AlphaSim correction with a voxel - level P<0.01 and a corrected threshold of P<0.05 was used.

In the perimenopausal group, correlation analyses were conducted between the function connectivity (FC) values of the DAN, GMV values, MRS scores, PHQ - 9 scores, the accuracy rate and reaction time of the Stroop color - word test, and sex hormone levels. After group analysis, the regions with significant FC changes between the two groups were identified, and the mean FC of each region in the perimenopausal group was extracted. Spearman partial correlation analyses were performed to evaluate the relationships between the mean FC values, GMV values of these regions, MRS score, PHQ - 9 score, the accuracy rate and reaction time of STROOP color - word, and sex hormone levels, with age and years of education considered as nuisance covariates.

### Sensitivity Analysis

To further assess the stability of our results, we conducted sensitivity analyses. We defined extreme values for age, follicle - stimulating hormone (FSH), estradiol (E2), and other key hormone levels as those beyond 2 standard deviations from the mean. Separate subsets of data were created by excluding participants with extreme values in age and hormone levels, respectively. Then, we re - performed all the statistical analyses, including the independent two - sample t - test for demographic and clinical data comparison, the general linear model analysis for DAN functional connectivity differences, and the Spearman partial correlation analysis for exploring relationships between variables.

### Exploratory Analysis

We employed exploratory data analysis methods to uncover potential patterns and relationships in the data. Specifically, we used hierarchical clustering analysis to group perimenopausal women based on their DAN functional connectivity, cognitive function scores (including STROOP test results), and hormone levels (FSH, E2, etc.). The Euclidean distance was used as the similarity measure, and the Ward’s method was applied for clustering.

## Results

### Demographic and Clinical Data

The demographic characteristics, scale, STROOP assessments, and sex hormone data for the two groups are presented in Table 1. There were significant differences in age, MRS score, PRL, FSH, E2, and the reaction time of the STROOP color - word test (P<0.05). No significant differences were found in years of education, PHQ - 9 scores, T, P, LH, and the accuracy rate of STROOP color - word between the two groups (P>0.05).

**Table 1.** Demographic, sex hormone levels and behavioral data between perimenopausal and premenopausal groups.

|  | Perimenopausal<br>group (n=25) | Premenopausal<br>group (n=25) | P value |
| --- | --- | --- | --- |
| Age (years) | 53.19±3.82 | 47.67±3.48 | <0.001 |
| Educations<br>(years) | 13.00±4.58 | 15.07±4.20 | 0.201 |
| PHQ-9 score | 2.38±1.63 | 3.20±3.60 | 0.409 |
| MRS score | 18.50±1.55 | 10.93±2.84 | <0.001 |
| PRL(ng/ml) | 12.05±8.60 | 19.01±5.88 | 0.014 |
| FSH(IU/L) | 24.25±10.58 | 8.07±3.78 | <0.001 |
| E2(pg/mol) | 22.84±11.00 | 101.47±70.34 | <0.001 |
| T(ng/dl) | 35.56±10.27 | 30.84±10.33 | 0.303 |
| P(ng/ml) | 0.17±0.18 | 0.31±0.34 | 0.161 |
| LH(mIU/ml) | 28.00±26.52 | 14.82±12.96 | 0.093 |
| The reaction time<br>of STROOP<br>(ms) | 1354.75±261.17 | 1094.37±146.16 | 0.002 |
| The accuracy<br>rate of STROOP<br>(%) | 97.88±3.81 | 96.73±4.38 | 0.097 |

PHQ-9, Patient Health Questionnaire-9; MRS, Menopause Rating Scale; PRL, prolactin; FSH, follicle stimulating hormone; E2, estradiol; T, testosterone; P, progesterone; LH, luteotropic hormone.

### Comparison of FC in the Resting - State DAN

The results of comparisons of brain regions with different FC in the DAN between the perimenopausal and premenopausal groups are shown in Table 2. After AlphaSim adjustment, with clusters ≥ 74 and P<0.05, the brain regions with enhanced FC in the DAN of perimenopausal women compared to premenopausal women included the right inferior parietal lobule (IPL) and the right angular gyrus (AG) (Fig. 1). The FC values of these two brain regions were significantly higher in the perimenopausal group than in the premenopausal group (Table 3, P<0.05).

**Figure 1.**
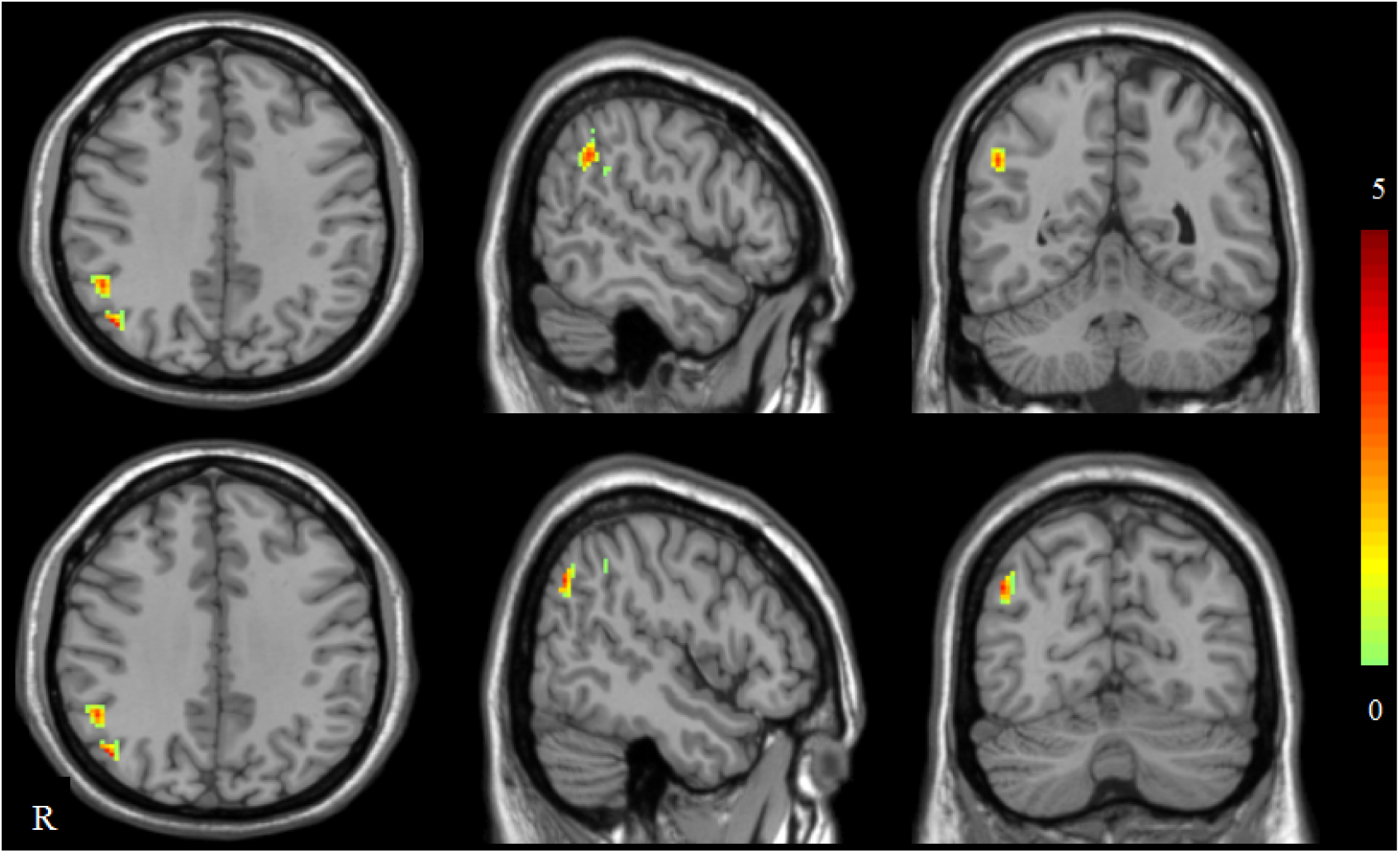
Brain regions with enhancing functional connection within DAN between the perimenopausal and premenopausal groups DAN, dorsal attention network.

**Table 2.** Brain regions with different functional connections in the DAN between the perimenopausal and premenopausal groups.

| Brain regions |  |  | BA | Cluster<br>size(voxel) | MINI<br>Coordination<br>(x,y,z) | Peak t score |
| --- | --- | --- | --- | --- | --- | --- |
| Right inferior parietal lobule |  |  | 40 | 83 | 52,-52,36 | 3.72 |
| Right angular gyrus |  |  | 39 | 82 | 48,-68,36 | 3.89 |

DAN, dorsal attention network; BA, Brodmann area; MNI, Montreal
Neurological Institute coordinate system or template; x, y, z, coordinates of primary peak locations in the MNI space.

**Table 3.**
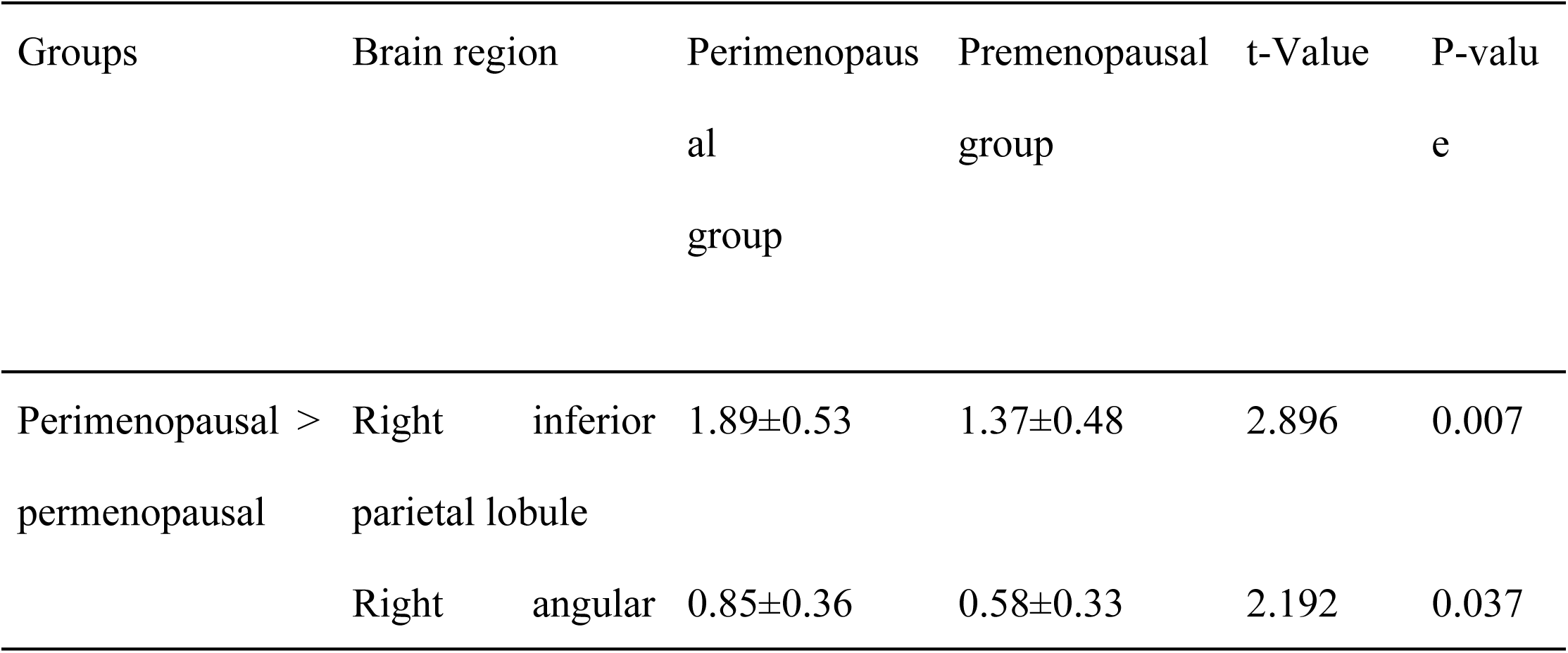
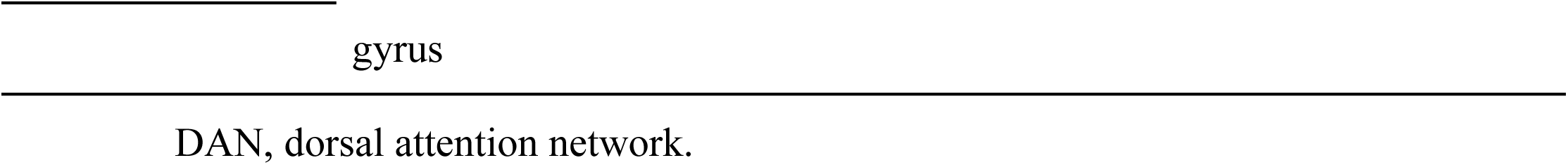
Functional connection values differences in DAN between the perimenopausal and premenopausal groups.

### Comparison of GMV in the Resting - State DAN

GMV values were extracted for the two brain regions (right IPL and right AG). There were no significant differences in GMV values of DAN between the perimenopausal and premenopausal groups (P > 0.05).

### Correlation Analysis

Spearman partial correlation analysis in the perimenopausal group showed that the FC value of the right inferior parietal lobule was significantly negatively correlated with the E2 level (P = 0.003, correlation coefficient: - 0.585) and positively correlated with the reaction time of STROOP (P = 0.001, correlation coefficient: 0.636; Fig. 2). The GMV values of the two ROIs in DAN showed no significant correlations with sex hormone levels, scales, and STROOP data.

**Figure 2.**
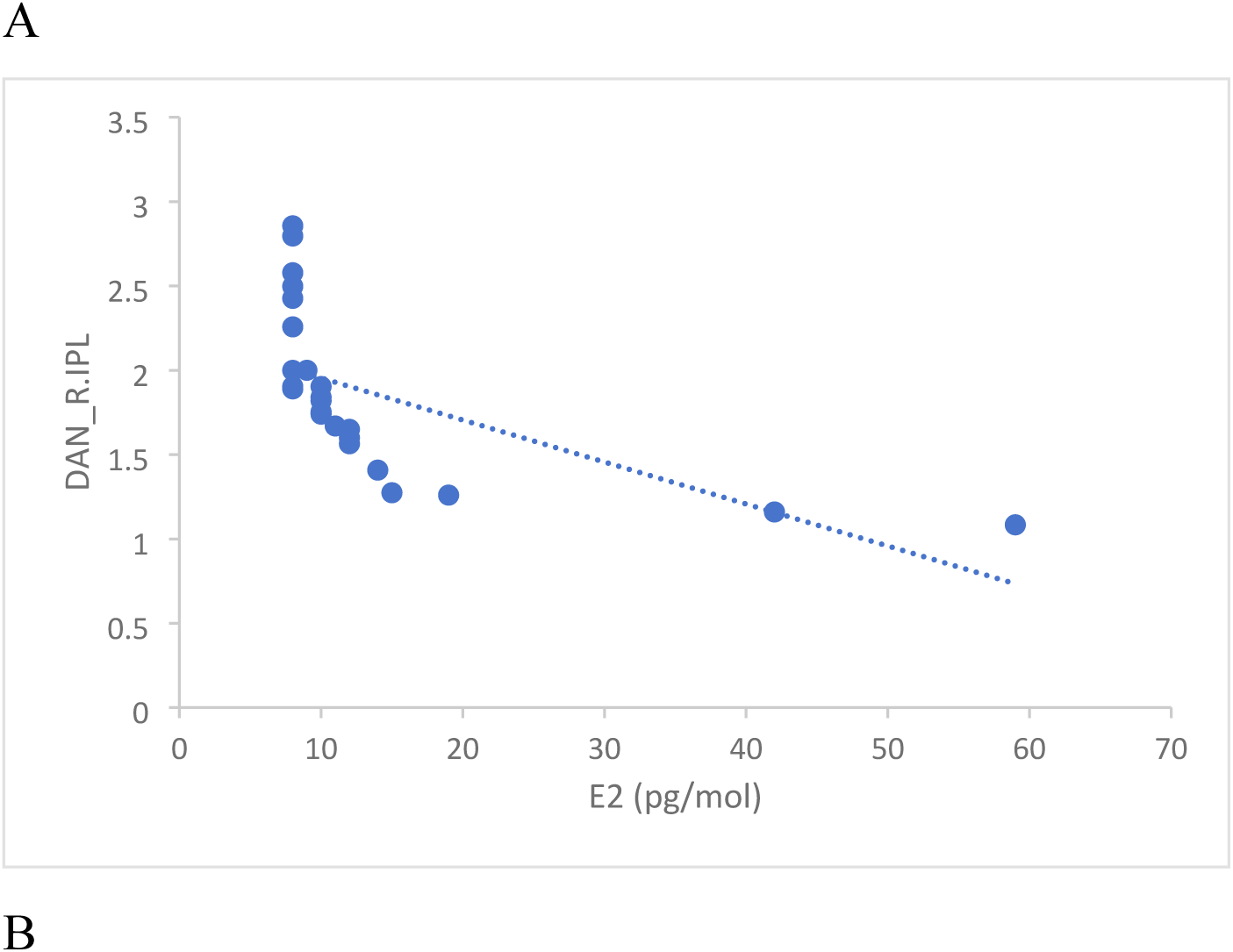
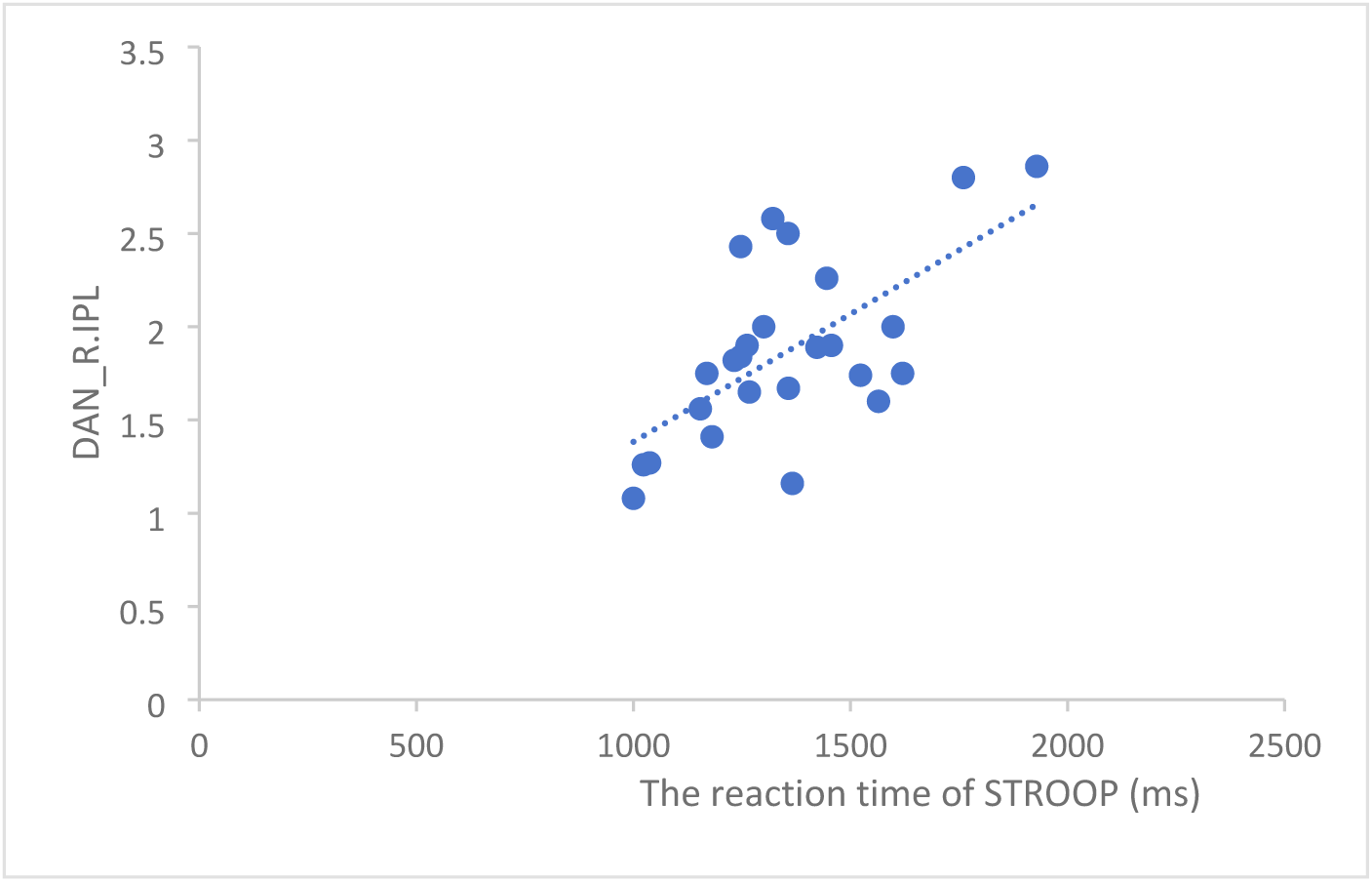
Correlation analysis results between functional connection values and clinical information in perimenopausal group. (A) Scatter plots between functional connection value of the right IPL in DAN and E2 in perimenopausal women. (B) Scatter plots between functional connection value of the right IPL in DAN and the reaction time of STROOP in perimenopausal women. R.IPL, right inferior parietal lobule; DAN, dorsal attention network; E2, estradiol.

Moreover, Spearman’s partial correlation analysis indicated no significant correlations between the GMV values of the two ROIs in DAN and age in both the perimenopausal and premenopausal groups. There were no significant correlations between age and sex hormone levels in the two groups.

### Sensitivity Analysis Results

After performing the sensitivity analyses, the main findings remained consistent. In the comparison of DAN functional connectivity, the enhanced functional connections in the right inferior parietal lobule (IPL) and the right angular gyrus (AG) in perimenopausal women compared to premenopausal women were still significant. The right IPL showed a significant increase in functional connectivity (P = 0.006) and the right AG also had a significant difference (P = 0.038). Regarding the correlation analysis in perimenopausal women, the negative correlation between the right IPL functional connectivity and E2 level persisted (P = 0.004, correlation coefficient: - 0.575), and the positive correlation with the reaction time of the STROOP test also remained significant (P = 0.002, correlation coefficient: 0.628). These results indicate that our original findings are robust to the influence of extreme values in the current dataset.

### Exploratory Analysis Results

The clustering analysis identified two distinct subgroups among perimenopausal women. Group 1 had relatively higher DAN functional connectivity in the right IPL and AG, better cognitive function (shorter reaction time in the STROOP test), and higher E2 levels compared to Group 2. In Group 1, the mean right IPL functional connectivity value was 2.05 ± 0.48, while in Group 2 it was 1.62 ± 0.51 (P = 0.02). The mean E2 level in Group 1 was 30.56 ± 10.23 pg/mol, significantly higher than that in Group 2 (20.12 ± 8.56 pg/mol, P = 0.005). These differences suggest that there might be different phenotypes within the perimenopausal population, which could be further investigated in future large - scale studies.

## Discussion

The perimenopausal period is a complex stage in a woman’s life, with significant hormonal and physiological changes. Our study aimed to investigate the functional changes of the DAN in perimenopausal women and their relationship with sex hormones and cognitive function.

The enhanced functional connectivity in the right IPL and right AG of the DAN in perimenopausal women is an interesting finding. The IPL is involved in various cognitive functions, such as attention, memory, and action - related functions^21–22^. The positive correlation between the right IPL functional connectivity and the reaction time of the STROOP test in perimenopausal women may suggest a decline in cognitive processing speed. However, this enhanced connectivity could also be a compensatory mechanism. As estrogen levels decrease during perimenopause, the brain may increase the connectivity in the IPL to maintain cognitive function ^23^.

The right AG is a crucial region for semantic processing, memory retrieval, and attentional control^24–25^. The increased functional connectivity in the right AG of perimenopausal women may reflect an attempt to cope with the cognitive challenges associated with hormonal changes. The lack of significant differences in cognitive task accuracy between perimenopausal and premenopausal women, despite the changes in DAN connectivity, further supports the idea of compensatory mechanisms in the brain.

The negative correlation between the right IPL functional connectivity and E2 levels in perimenopausal women is consistent with previous studies. Estrogen plays a significant role in maintaining brain function. When E2 levels decline, the DAN may need to enhance its connectivity to compensate for the potential negative effects on cognitive function^26–27^. The results of our sensitivity analysis are reassuring, indicating that our main findings are not unduly influenced by extreme values in the data. This provides additional confidence in the reliability of our results. Sensitivity analysis is a widely - used approach to assess the stability of research findings. As described in Tabachnick and Fidell’s “Using Multivariate Statistics” ^28^, this method helps to identify the influence of outliers on the overall results. However, it should be noted that sensitivity analysis has its limitations and cannot completely rule out the impact of other unmeasured factors.

The exploratory analysis identified potential subgroups within perimenopausal women based on DAN functional connectivity, cognitive function, and hormone levels. This finding suggests that there is heterogeneity within the perimenopausal population. The hierarchical clustering analysis used in our study follows the principles and methods described in Everitt et al.’s “Cluster Analysis”^29^. Future large - scale studies are needed to further explore these subgroups, understand the underlying mechanisms, and determine whether different subgroups require different treatment strategies.

When comparing our results with existing studies, we found both similarities and differences. A recent large - scale study^8,14,30^ also reported changes in DAN connectivity during perimenopause, which is consistent with our findings and validates our results to some extent. However, they also detected changes in other brain regions that we did not observe. This may be due to differences in sample size, research methods, or participant characteristics. Their larger sample size might have allowed for the detection of more subtle effects. In contrast, our study focused on a more in - depth exploration of the relationship between DAN, sex hormones, and cognitive function, providing unique insights into this complex relationship.

However, this study has several limitations. Despite the robustness shown in the sensitivity analysis, the relatively small sample size remains a major limitation. A larger sample size would increase statistical power, enabling the detection of smaller effects and improving the generalizability of our findings. Additionally, the cross - sectional design of this study does not allow us to establish causal relationships between hormonal changes, DAN functional connectivity, and cognitive function. Longitudinal studies are required to better understand the dynamic changes during the perimenopausal period.

## Conclusion

This study used resting - state fMRI to investigate the differences in the DAN between premenopausal and perimenopausal women. The results showed that cognitive function was associated with enhanced DAN connectivity in perimenopausal women, especially in the right IPL and right AG. Correlation analyses revealed relationships between estrogen levels, cognitive function, and DAN. Sensitivity and exploratory analyses provided additional insights into the data, and comparison with existing studies enhanced the understanding of our findings. These findings contribute to our understanding of the neural mechanisms underlying cognitive changes during perimenopause and may provide insights for the diagnosis and treatment of perimenopausal cognitive dysfunction.

However, due to the limitations of this study, future research with larger sample sizes and longitudinal designs is needed to further explore these relationships.

## Data Availability

All relevant data are within the manuscript and its Supporting Information files.

## Acknowledgments

The authors are deeply grateful to all participants involved in this study and to all doctors and researchers who contributed to the research.

## Authors’ Contributions

LNN and LHJ designed the study, ZY and FWQ collected data, LNN and ZY wrote the manuscript, and LNN and LHJ revised the manuscript. All authors read and approved the final manuscript.

## Ethical Statement

This study was approved by the Ethics Committee of the Second Hospital of Tianjin Medical University. All participants provided written informed consent, and the study was conducted in accordance with the Declaration of Helsinki.

## Data Availability

The datasets used and/or analyzed during the current study are available from the corresponding author upon reasonable request.

